# Modern contraceptive use among adolescent girls and young women in Benin: a mixed-methods study

**DOI:** 10.1101/2021.06.07.21257758

**Authors:** Noudéhouénou Crédo Adelphe Ahissou, Lenka Benova, Thérèse Delvaux, Charlotte Gryseels, Jean-Paul Dossou, Sourou Goufodji, Lydie Kanhonou, Christelle Boyi, Armelle Vigan, Koen Peeters, Miho Sato, Mitsuaki Matsui

## Abstract

**Objectives:** The study aimed to assess the determinants of modern contraceptive method use among young women in Benin.

**Design:** A mixed-methods design.

**Setting and participants:** We used the Benin 2017-18 Demographic and Health Survey datasets for quantitative analysis. Data collection was conducted using multiple-cluster sampling method and through household survey. Qualitative part was conducted in the city of Allada, one of the Fon cultural capitals. The participants were purposively selected.

**Outcomes:** Contraceptive prevalence rate, unmet need for modern method, and percentage of demand satisfied by a modern method for currently married and sexually active unmarried women were measured in the quantitative part. Access barriers and utilization of modern methods were assessed in the qualitative part.

**Results:** Overall, 8.5% (95%CI: 7.7-9.5%) among young women ages 15 to 24 were using modern contraceptives and 13% (95%CI: 12.1-14.0%) among women ages 25 or more. Women 15-24 had a higher unmet need, and a lower demand satisfied by modern contraceptive methods compared to women aged 25 or more. 60.8% (56.9-64.7%) of all unmarried young women had unmet need for modern contraceptives. Young women were more likely to use male condoms which they obtain mainly from for-profit outlets, pharmacies, and relatives. The factors associated with demand satisfied by a modern method were literacy, being unmarried, knowing a greater number of modern contraceptive methods, and experiencing barriers in access to health services. On the other hand, the qualitative study found that barriers to using modern methods include community norms about pre-marital sexual intercourse, perceptions about young women’s fertility, spousal consent, and the use of non-modern contraceptives.

**Conclusion:** Contraceptive use is low among young women in Benin. The use of modern contraceptives is influenced by socio-demographic factors and social norms. Appropriate interventions might promote comprehensive sexuality education, increase community engagement, provide youth-friendly services, and address gender inequalities.

**Strengths and limitations of this study:**

- This study used a nationally representative survey data which allows generalizability at the national level.
- We used a mixed methods design which enables us to compare and discuss the large-scale quantitative data with qualitative information from the ground.
- The quantitative information could have been affected by recall errors and biases.

## INTRODUCTION

Despite efforts toward making access and use of contraceptive services a basic reproductive right for all women, many countries still face high rates of unintended and unwanted pregnancies,[1, 2]. In sub-Saharan Africa alone, about 14 million unintended (unwanted or mistimed) pregnancies occur every year,[3]; and adolescent girls and young women 15 to 24 years old are the most vulnerable group,[4]. The reasons behind this situation include the high prevalence (about 70%) of sexually active young women with low utilization of effective contraceptive methods (less than 10%),[4]. Additionally, unmarried sexually active adolescents are likely to have a high unmet need for contraception, which increases their risk of unintended pregnancies,[5, 6]. In developing countries, nearly half (49%) of pregnancies are unintended among adolescent girls of 15 to 19 years old,[7]. Adolescence is viewed as the starting point in the continuum of care for reproductive, maternal, neonatal, and child health; and is a phase when poor access and utilization of contraception are likely to result in poor health outcomes across the continuum of care,[8, 9]. Early and unintended pregnancies result in increased risks of maternal mortality and morbidity, premature births, low birth weight, unsafe abortions, and social consequences such as stigmatization, school dropout, and poverty,[10, 11].

In Benin, although the use of modern contraceptives has been slowly increasing since 2006, it is still relatively low. The modern contraceptive prevalence among all women reached 12% in 2018 compared to 6% in 2006,[12]. At the same time, almost half (48%) of all adolescent girls age 15 to 19 are sexually active, and one in five girls has already had a child or is pregnant,[12]. According to the government of Benin (2017), only 5.4% of women ages 15 to 24 were using modern contraceptive methods in 2017,[13]. Recent data showed a total fertility rate of 5.7 among all women of reproductive ages 15 to 49 years old, and the modern contraceptive prevalence rate (mCPR) was estimated at 12%,[12]. Of all pregnancies in the country, 19% were unintended,[14]; and in 2017, both the maternal mortality ratio and infant mortality rate remained high at 397 per 100,000 live births, and 30 per 1,000 live births, respectively,[6, 12].

Existing studies on the use of modern contraceptive methods in Benin largely reported on women of reproductive age as a whole, rather than focusing on specific age groups, [15-20]. MacQuarrie suggested that young women should be studied separately, as they do not have the same needs for or access to contraception as adult women,[21]. However, for Benin, the literature is sparse on the use of reproductive health services among adolescent girls and young women. <u>There two studies which focused on young women’s contraceptive use in African countries including Benin,[22, 23]. However, these studies employed quantitative method</u> and did not explore in-depth factors that affect decision-making such as adolescent girls and young women’ s perceptions, preferences, and interactions with peers and providers. <u>Moreover, little is known about the contraceptive practices and needs of sexually active and inactive young women specifically.</u>

Therefore, this study aimed to assess contraceptive use, perceptions, and practices among adolescent girls and young women in Benin. Specifically, we used secondary quantitative data to examine the rates of modern method use, unmet needs for modern contraception, and to evaluate the association between various factors and the demand satisfied for a modern contraceptive among sexually active young women. We then used qualitative methods to further explore the socio-cultural factors influencing the use of modern contraceptive methods among young women of ages 18 to 24 years.

## METHODS

### Study design

The study was conducted using a sequential explanatory mixed-methods design where the collection and analysis of quantitative data were followed by the collection and analysis of qualitative data. The qualitative results thus aim to explain and interpret the findings of the quantitative study,[24].

### Quantitative strand

#### Data source

We used the most recent Benin Demographic and Health Survey (BDHS), conducted between November 2017 and February 2018 ,[12]. The BDHS2017-18 was a nationally representative cross-sectional survey conducted by the National Institute of Statistics and Economic Analysis and funded by the USAID. The BDHS 2017 - 18 employed a two-stage stratified cluster sampling during which data were collected with standardized questionnaires.

#### Study population

Included in the study were women ages 15 to 49 years at the time of the survey, residing in sampled households, and who agreed to respond to the BDHS 2017-18 . We defined adolescent girls as those 15 to 19 years old and young women as those 20 to 24 years. In descriptive analyses, we compared indicators of contraceptive use in each of the age groups to all women 25 years and older. In the analysis of the determinants of modern contraceptive methods use, we focused on women who reported to be sexually active (married and unmarried) because they are exposed to the risk of pregnancy, and we used the combined age groups 15 to 24 years due to sample size considerations.

#### Definitions

Some definitions of indicators from the BDHS 2017-18 were adopted, while others were generated or recategorized as necessary for our study-specific objectives.

#### Modern method

Respondents were asked which method of contraception they were using at the time of the survey. If they selected more than one method (for example, implant and condom), only the most effective method was noted in the dataset. Based on the definition of Hubacher and Trussell (2015), we considered modern contraceptive methods to include: oral contraceptive pills, intrauterine devices (IUD), injectables, diaphragm, male/female condoms, female/male sterilization, implants/Norplant, foam, or jelly, and emergency contraception,[25]. All other methods were grouped as traditional methods.

##### Current modern method users

Women who reported using any modern contraceptive method at the time of the survey were considered modern method users.

##### Contraceptive prevalence rate

The proportion of women who reported using a method at the time of the survey. This was estimated for modern contraceptive methods among different groups of women based on their marital status.

##### Sexual activity

Women who reported sexual intercourse 30 days preceding the survey were categorized as sexually active.

##### Source of modern method

The source of the modern method is the most recent source of provision among current modern methods users. We grouped sources of modern methods into public sources (government hospital, government health center, mobile clinic, government community health worker, and other public sectors), non-public sources (private hospital/clinic, private clinic or doctor, and private mobile clinic), for-profit outlets (pharmacy, family planning clinic, and shop), relatives and other sources (friends/relatives). A second categorization in the analysis was done based on the theoretical capacity model of providers proposed by Radovich et al.,[22]. It divides the providers into two groups, ‘ comprehensive’ or ‘limited’. Comprehensive providers are public or private providers that can provide both short- and long-acting methods. Limited providers include service locations, for-profit outlets, partners, and friends, which do not generally provide long-acting methods.

##### Unmet need for modern methods

Unmet need for contraception describes women who are at risk of pregnancy (married or sexually active), do not wish to be pregnant, but are not using a modern contraceptive method. We applied an algorithm proposed by Bradley et al. (2012) to identify ‘unmet need for limiting’ and ‘ unmet need for spacing’ among married women and sexually active unmarried women,[26]. Also, we adopted the definition of FP2020 (2021) to compute the ‘unmet need for modern methods’,[4]. Because women using a traditional method are less protected against pregnancy, they were included with the non-users and considered to have an unmet need for modern contraception,[27]. Unmet need was estimated among the denominator of all women regardless of whether they were in need of family planning or not.

##### Percentage of demand satisfied by a modern method

This is the percentage of women at risk of pregnancy and who do not wish to become pregnant, who are using modern methods. The numerator is current modern method users, and the denominator is all women in need of family planning (current users of any contraceptive methods and women with unmet need for family planning).

##### Existence of barriers to access to health services

Women were asked whether getting permission to go, getting money to pay for services, the distance to travel, or going alone to a health center were barriers in their access to health services. We created a binary variable categorizing women who said that they had none of these issues versus those who had one or more.

##### Core indicators

The core indicators include ‘contraceptive prevalence rate’, ‘unmet need for modern method’, and ‘percentage of demand satisfied by a modern method’ for currently married and sexually active unmarried women. Demand satisfied by a modern method among women ages 15 to 24 in need of family planning was the main outcome variable in the logistic regression.

##### Explanatory variables

We included in the logistic regression the respondents’ socio-demographic characteristics (literacy, ethnicity, religion, marital status [married/cohabiting at the time of the survey or no], parity, and household wealth quintile); knowledge of modern contraceptive methods; and the existence of barriers to access health services.

#### Data analysis

Data analysis was carried out using Stata (version 13, Stata Corp., Texas, USA). We conducted a descriptive analysis to describe the socio-demographic characteristics of the study population within three age groups (15 to 19, 20 to 24 , 25 or older). We used logistic regression to conduct bivariate and multivariable analyses to assess the associations between the explanatory variables and modern method use among sexually active women of ages 15 to 24 years in need of contraception. We retained all explanatory variables in multivariable logistic regression. We used the *svyset* command to adjust for clustering, stratification, and the sampling weights of individual women in all analyses.

#### Missing data

There was no missing data in any variables used except for the source of the modern method. Of condom users 15 to 49 years old, 26 out of 1805 (1.4 %) had a missing source of method; and 19 of them were 15 to 24 years old. We grouped them within the “ pharmacy” category, which is the most common source for condom users.

### Qualitative strand

#### Study site and population

Data collection was conducted in the city of Allada, one of the Fon cultural capitals – the largest ethnic group in Benin - and located in southern Benin about 50 km from Cotonou, the largest city of Benin. The region offers the advantage of being semi-rural with diverse religions and care practices such as spiritual care, alternative care, and biomedicine,[28]. Spiritual healing involves using mystical practices, advice prescribed by traditional healers which are viewed as effective in treating health problems. Such practices could include consulting deities, performing rituals, using leaves, or consuming herbal teas. Alternative care refers to the use of herbal infusions and powders, as well as the use of certain leaves and barks to address health issues.

We restricted the target population of the qualitative study to women ages 18 to 24 years old due to consent issues with minors. We collected data from two main groups: young pregnant women from whom we wanted to learn about access barriers and utilization of modern methods based on their experiences, particularly in cases where pregnancy might be unintended. They were recruited after pre or postnatal visits, or consultation about family planning services in the main public health center of Allada.

Healthcare providers helped us identify women that met our inclusion criteria (pregnant young women). We later approached them face- to- face outside the health facilities after consultations. Non-pregnant adolescent girls and young women were recruited in schools through purposive sampling, and in the community using convenience and snowball sampling techniques. In the community, we visited public places (markets, school yards, parks) and approached young women face- to-face and were selected if they met our inclusion criteria and were willing to participate in the study. New potential participants were then referred by previous informants.

Since the use of modern contraceptive methods may also be affected by other actors in decision-making and service provision, we collected data from young men ages 18 to 24, health care providers in public and private local health centers, relatives of adolescent girls, and the leaders of a non-governmental organization (NGO) which promotes contraceptive methods in the region. Those informants were all recruited by purposive sampling techniques.

#### Data collection

Data were collected through in-depth interviews and focus group discussions with young people, females, and males separately. The group discussions were conducted in isolated public places (parks, youth gathering places) to ensure quietness and that participants speak with ease and without fear of being seen by relatives. We conducted the discussions mainly in the Fon language. The number and characteristics of the participants are shown in Table 1.

**Table 1.**
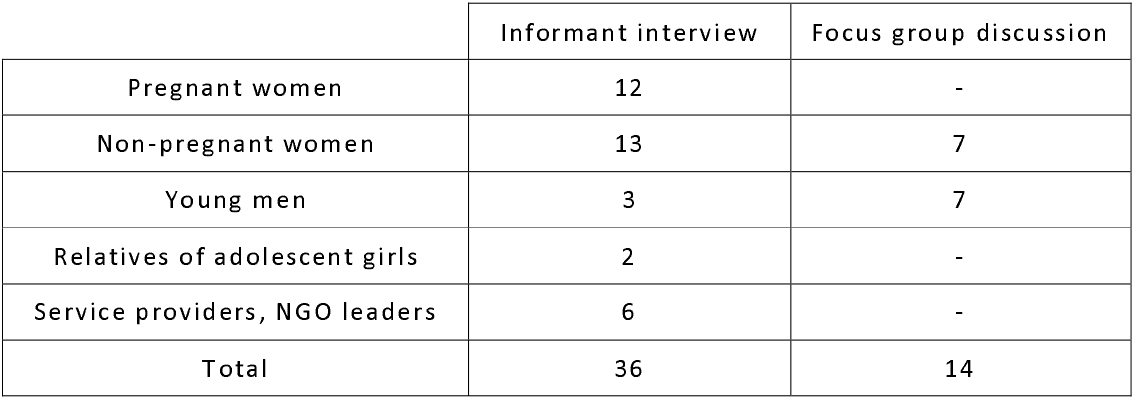
Numbers and characteristics of participants in informant interviews and focus group discussions.

Based on an adapted theoretical framework from the PASS Health Seeking Behaviour model of Hausmann-Muela et al.,[29], and the first findings from the quantitative analysis, we designed interview guides specific to each target group. The question guides focused on perceptions of ideal family size, knowledge of contraceptive methods, and decision-making for contraceptive use.

#### Data analysis

All recorded sessions were transcribed into French. Those that were not conducted in French were translated. A content analysis of transcripts was done using the software MAXQDA 11. In the inductive analysis, narrative data was parsed and categorized into units of meaningful information and the categories were linked to understanding young women’ s decision-making for contraception.

## RESULTS

### Quantitative strand

Overall, 15,928 women of reproductive age (15 to 49 years old) were surveyed on the DHS; 39.4% (n=6,251) were 15 to 24 years old. The sociodemographic characteristics of the respondents by three age groups (15 to 19 years old; 20 to 24 years old; and 25 years or older) are shown in Table 2 .

**table 2.**
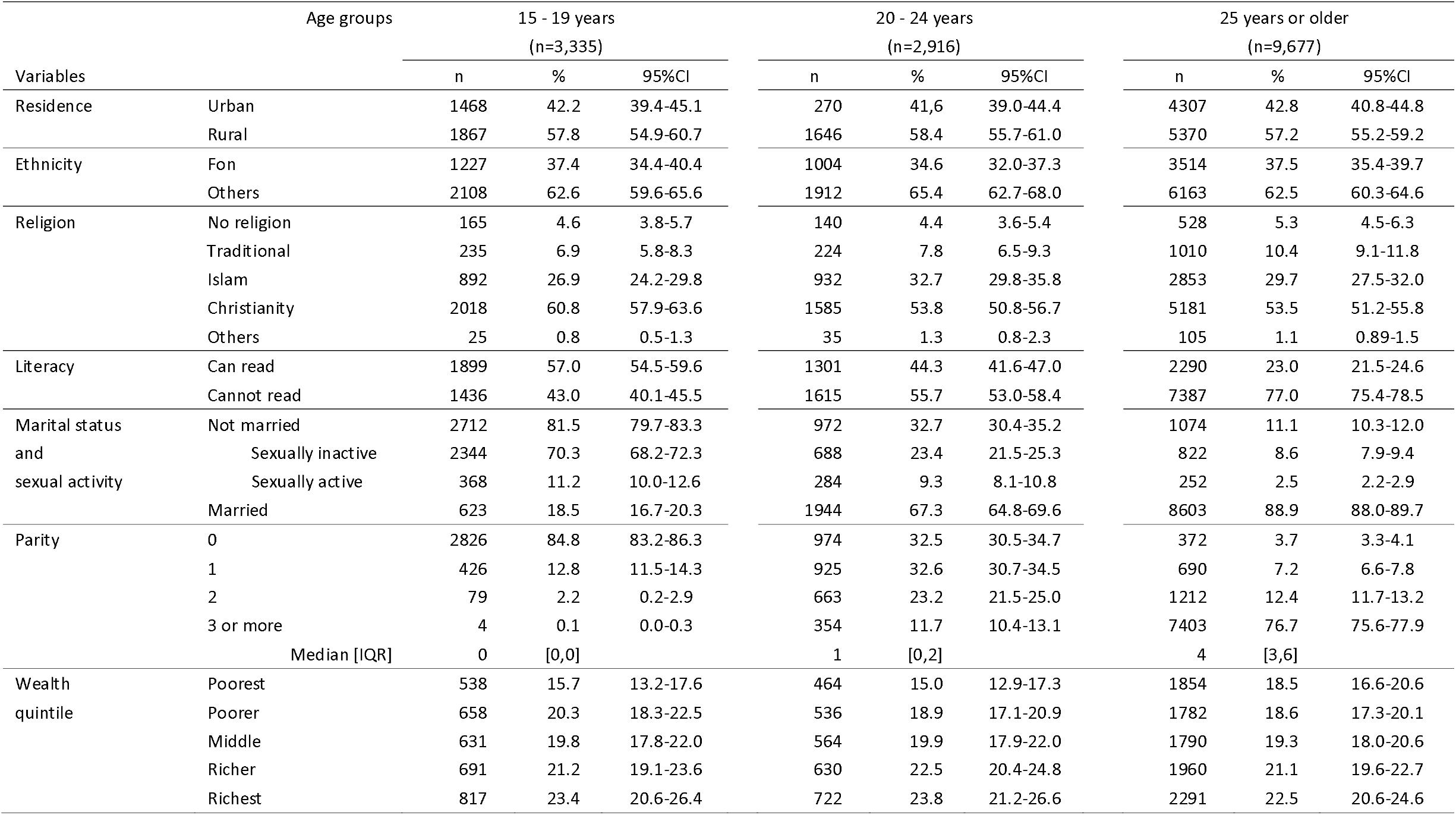
Socio-demographic characteristics of the quantitative analysis sample (Benin DHS 2017-18); N=15,928 (women aged 15-49)

Modern contraceptive use was 11.2% (95%CI: 10.6-12.0%) among all women of reproductive age (15 to 49 years old) versus 8.5% (95%CI: 7.7-9.5%) among women aged 15 to 24, and 13.0% (95%CI: 12.1-14.0%) among 25 years old women or older.

Contraceptive practices and knowledge in the study population are shown in Table 3 , where we compared four groups: adolescents and young women (15-24 years) and women 25 years or older, (separately married and unmarried sexually active). Among all women, the unmet need for modern contraception was significantly higher among young unmarried sexually active women (60.8%) compared to all other groups. Specifically, among women in need of family planning, the percentage of demand satisfied by a modern method was lowest among 15 to 24 years old married women (17.3%).

**Table 3.**
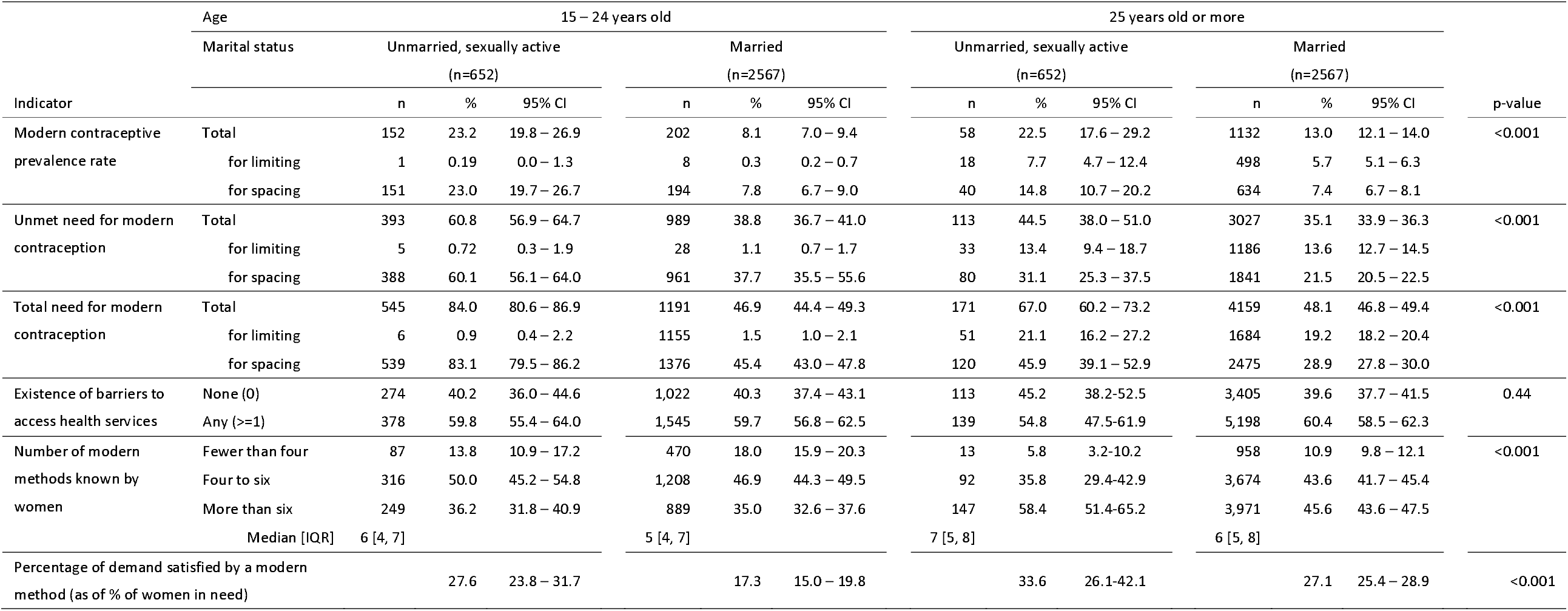
Need for, demand for, and knowledge of modern contraception among sexually active women in Benin (n=12,074)

Table 4 shows the method mix among users of modern 281 methods. Among unmarried sexually a ctive women ages 1 5 to 2 4 years, 6 3.4% of modern c ontraceptive users were using short-term methods, and more than 60% of these being male condom users. The use of modern contraceptives was lower among married young women (27.1%), sexually active unmarried women ages 25 or above (25.7%), and among married women 25 years old or more (19.3%). Although there was no substantial dif ference in long-term method use among married women (72.9% vs 80.7%) of 15 to 24 years old and 25 or older, the use of implants and IUDs was lower among young women (49.4% v s 6 1.0%). R egarding t he source of c ontraceptives, u nmarried sexually active young women obtained their contraceptives from for-profit outlets or relatives (62.1%) or comprehensive providers (36.2%) . The public sector was the main source of methods among young and adult married women (77.0% and 78.9%).

**Table 4:**
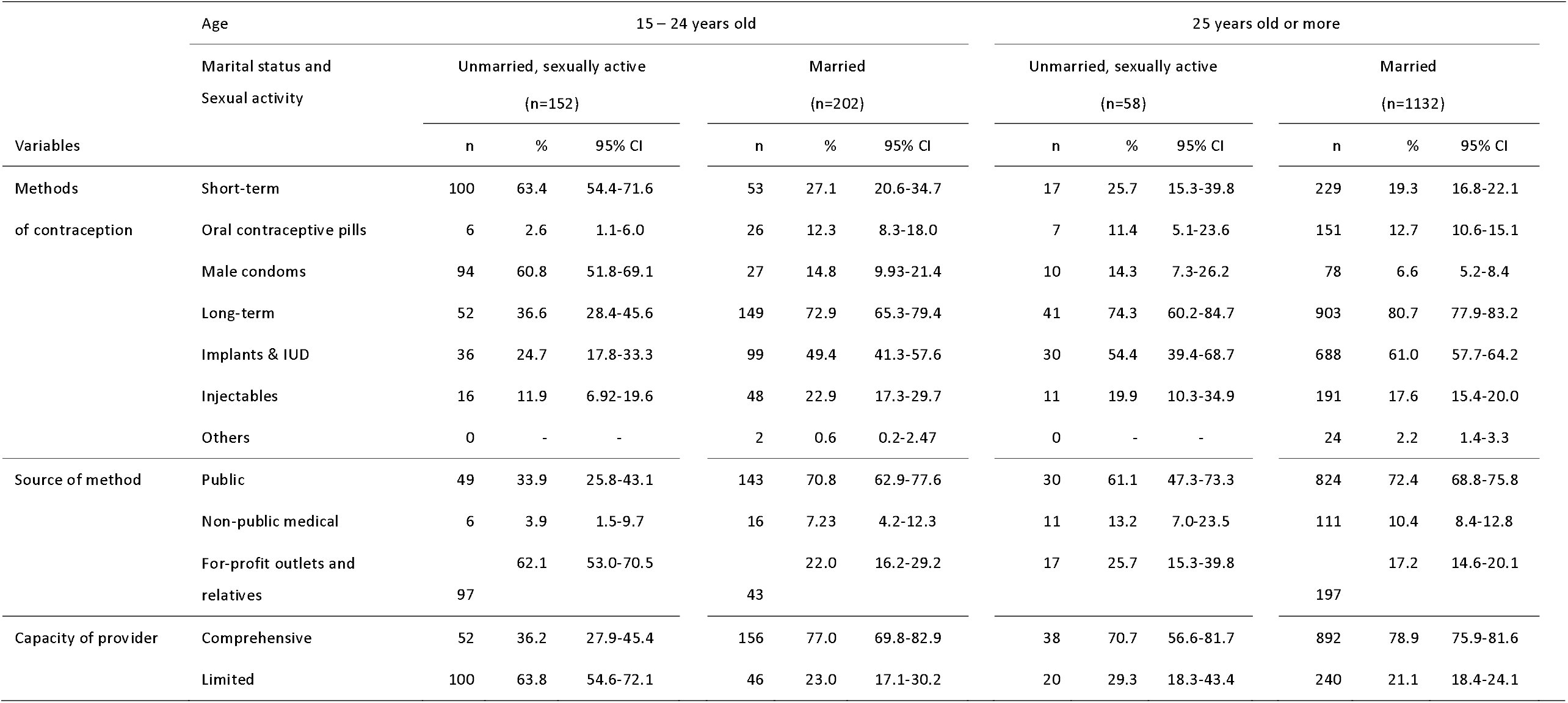
Modern contraceptive practices among unmarried and married sexually active modern contraceptive users, by age group (n=1544)

#### Factors associated with demand satisfied by a modern method

We examined the demand satisfied by a modern method among women ages 15 to 24 years in need of contraception (n=1736). Table 5 shows the percentages of modern method users and the crude and adjusted odds ratios of each factor on the use of modern methods. The multivariable analysis showed that being unmarried, higher parity, existence of barriers in access to health services, and a higher number of modern methods known by a woman were positively associated with demand satisfied by a modern method.

**Table 5.**
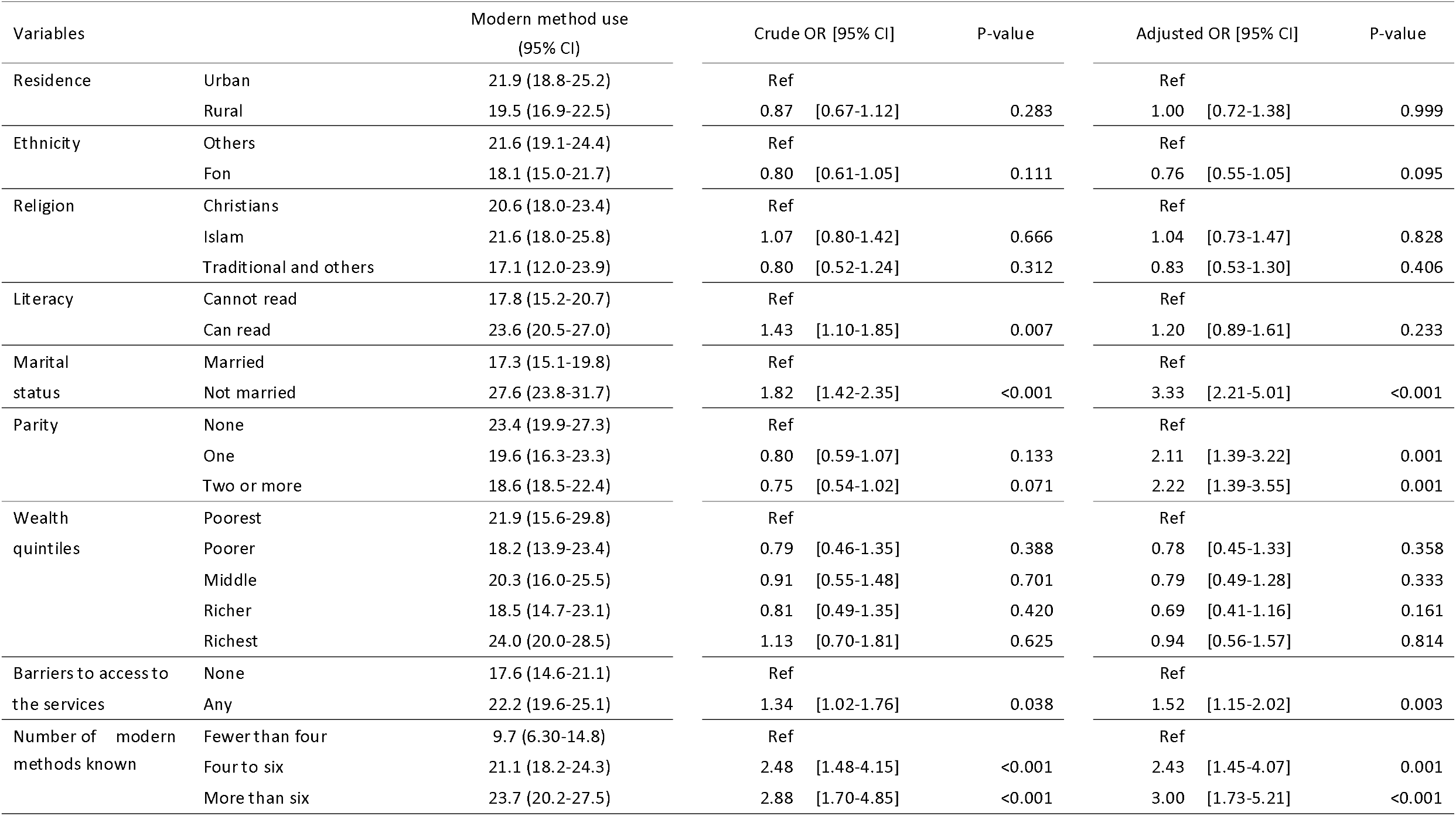
Percentages and factors associated with modern method use among 15-24-year-old women in need of contraception (n=1,736)

### Qualitative strand

We identified three main barriers in access to and utilization of contraception services among adolescents, which are (i) socio-cultural perceptions related to sexuality and fertility and (ii) perceptions about the purpose and effectiveness of contraception. A conceptual model of the barriers is shown in Figure 1.

**Figure 1.**
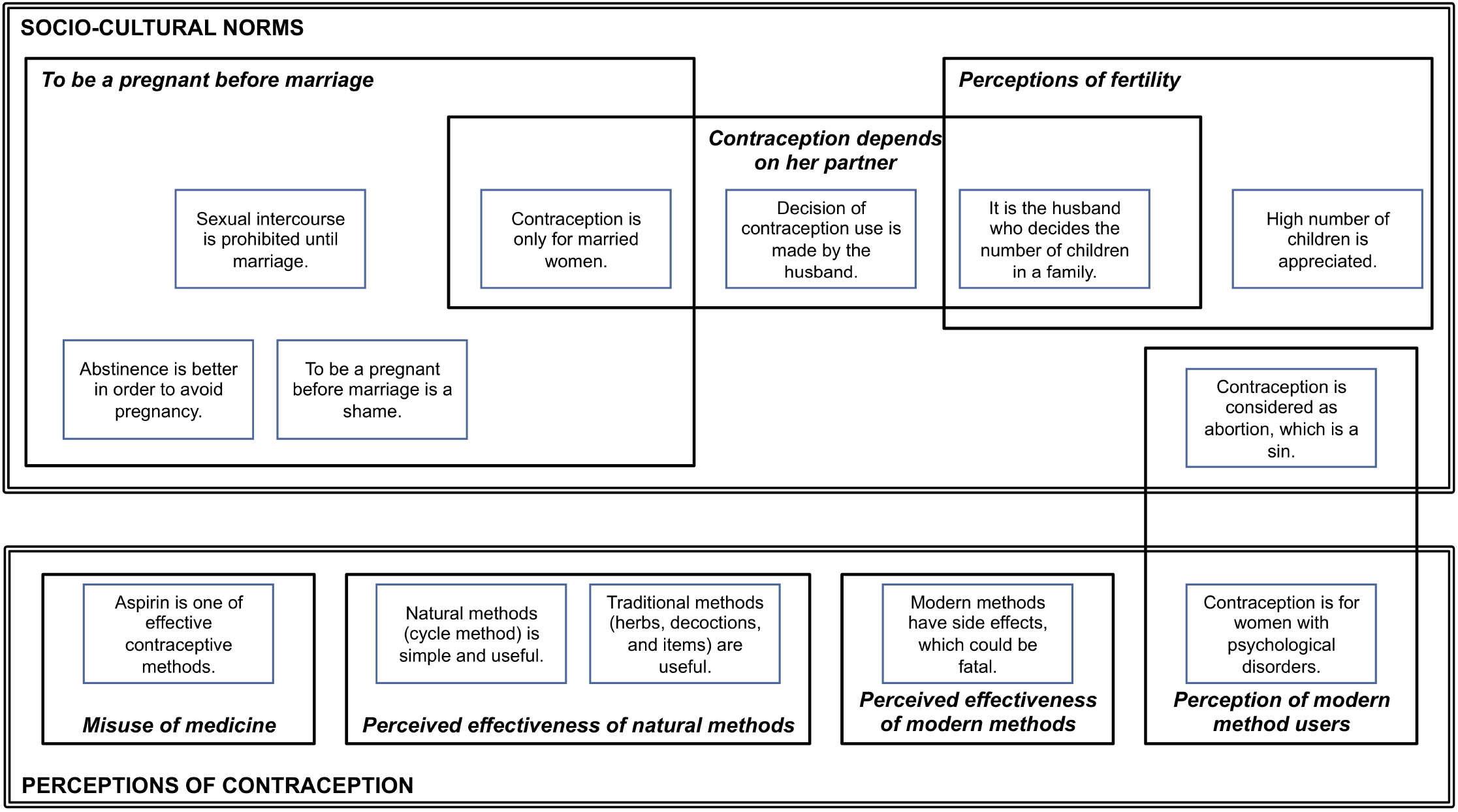
A conceptual model of the barriers to access and utilization of contraception services from qualitative analyses.

#### Sociocultural norms

Sociocultural norms including gender norms, cultural, and religious perceptions affect people’s perceptions about young women’s fertility and the need for contraception in Allada. These norms include views about pregnancy before marriage, male partners’ role in women’s contraceptive use, local perceptions of women’s fertility, resulting in different perceptions of contraception such as the misuse of biomedicine, perceived effectiveness of natural methods, mistrust in modern methods, and negative views about modern method users.

#### To be pregnant before marriage

There is a common negative perception about sexual intercourse among adolescents. Becoming pregnant as an adolescent and before marriage is not well viewed in the community as judged as promiscuity. Therefore, unmarried young women avoid such judgment through the use of traditional methods such as abstinence or fertility awareness. This allows young women to avoid exposing their pre-marital sexual activity publically. This situation prevents unmarried young women from using contraceptives.

> *“To avoid pregnancy, one must abstain from sex. If a girl really wants to avoid shame in front of her classmates, she must refrain from sex*.*” (less than 20 years old/pregnant)*

##### Perceptions regarding the ideal family size

In the study area, there was cultural pressure on having a certain number of children based on social norms about the ideal family size. It implies that when a young woman gets married, she may not be allowed to use contraception until she has several children.

> *“If we say that someone has enough children, it will be from seven. That is when the person will say, ‘Yes, I do have children*.*’” (less than 20 years old/male)*

None of the respondents mentioned cultural pressures that might force young unmarried women to have children early.

##### Decision-making dynamics

Women do not often have the decision-making power to use contraception.

> *“One day, some people went door-to-door to inform us of the presence of an NGO at the health center for family planning services. I then informed my husband and did the injection for 3 months. But after he forbade me to continue it, because of rumors about other women’s bad experiences. When your husband refuses you have no choice*.*” (20-24 years old pregnant)*

#### Perceptions of contraception

##### Contraception perceived as abortion

Religious beliefs, especially Catholicism, also affect the utilization of contraception.

> *“In my opinion, ‘planning’ is an abortion, a deadly sin. We are killing babies without knowing and it is against the ten commandments of God. We are also killing ourselves because we do not know the consequences of those things in our body*.*” (less than 20 years old/non-pregnant)*

#### Perceptions of modern method users

Respondents believed that the use of contraception facilitates sexual promiscuity. Contraception was also considered to be a eugenic procedure.

> *“When she was 12 years old, people told me to have her adopt a contraceptive method. I thought it was asking her to prostitute herself and I did not. Here now, as you see, she came home with pregnancy while she is still having her training*.*” (Relative of pregnant)*

#### Perceived effectiveness of natural methods

Faith in natural methods contributes to reluctance to use modern contraceptives among young women.

> *“I do not plan to use any of those things [modern methods]. My family members told me they will prepare an herbal tea for me and I will not get pregnant for the period I want - they know how to do it*.*” (less than 20 years old/pregnant)*

Beliefs were led by fear of side effects of modern methods, and these opinions were in part based on rumor or hearsay.

> *“ It happened to one of my relatives. She chose the one that is inserted in the arm. Three months later, she had no more periods. Then when she had menstrual periods again, it was too much. It was when it became excessive that she had to go where she had it inserted for removal. Finally, she died*.*” (20-24 years old/female)*

#### Misuse of biomedicine

Some also perceived ‘aspirin’ to be effective in preventing pregnancy and used it as a type of modern method.

> *“I often use Sedaspir [commercial name of acetylsalicylic acid] and it works well. I buy the box at the pharmacy for 1500 franc CFA [equivalent to 3 US dollars]*.*” (20-24 years old/pregnant)*.

## DISCUSSION

We investigated modern method use, unmet need for contraception, and the reasons why modern methods were not used; focusing on adolescent girls and young adult women (15 to 24 years old) using the most recent Benin Demographic and Health Survey (BDHS 2017-18) combined with a qualitative study conducted in the Allada area in Benin.

Our quantitative analysis revealed that characteristics of modern method use differed between unmarried sexually active and married young women ages 15 to 24 . Among all women, young unmarried sexually active women had both the highest modern contraceptive use and the highest unmet need for modern contraceptives. Modern method users tended to adopt more short-term than long-term methods, especially the male condoms which they acquired from providers with a limited range of methods. Young married women had the lowest percentage of demand satisfied by a modern method among women who wished to delay or limit births. Factors associated with modern method use among young women in need of contraception were being unmarried, having at least one child, experiencing barriers in access to health services, and knowing a greater number of modern contraceptives. As suggested by the qualitative analysis, the reasons underlying the high unmet need for modern contraceptives in young women are related to the access and use of modern methods which can be categorized as ‘ sociocultural norms’ and ‘ perceptions of contraception’.

Young unmarried women had the highest unmet need for modern contraceptives as a percentage of all women, which may explain the high rate of unintended pregnancy in Benin. Very few unmarried young women want to have a child due to community negative perceptions about pre-marital sexual intercourse and pregnancy, therefore in need of contraception. However, as unmarried young women are considered in the communities not to be ready for sexual intercourse they are reluctant to request contraceptives despite their wish to avoid a pregnancy,[19, 20]. Moreover, the fear of adverse effects contributes to young women’s non-use or discontinuation of modern contraceptives,[20, 30, 31].

Among unmarried young women in need of family planning, those with demand satisfied by a modern contraceptive tend to use male condoms, the most adopted modern method in Benin (45 .3%), which can be easily obtained from for-profit outlets, partners, and relatives. This finding is similar to that of Ali and Cleland,[32], secondary data analysis from 36 African countries which showed that male condoms constitute 51.3% of modern methods among unmarried women ages 15 to 24 years. Our qualitative analysis suggested that while access to male condoms has fewer barriers, male partners are in most cases the ones to obtain the method for use. Consequently, the decision to use a male condom is likely to be made by the male partner whose failure to acquire the method or unwillingness to use could be a barrier to the young woman’s contraceptive use. Male condoms are obtained in shops, pharmacies, or from non-governmental organizations during awareness-raising campaigns. Also, young women may assume that male partners obtain contraceptives mainly from pharmacies which could explain why pharmacies and relatives are the most frequent sources of methods reported in the DHS2017-18. Young people’s choice of providers including shops and pharmacies is based on their values for discretion, respect of confidentiality, and the quickness of the service delivery. This is in line with the findings of Murray et al.,[4], which suggested that the preference of pharmacies by young women in requesting contraceptive services may be driven by their perception that the providers are youth friendly.

Also, young married women in need of family planning had a lower percentage of demand satisfied by a modern contraceptive compared to unmarried young women and older married women. Most young women want to have a child once they get married which explains the low unmet need as a percentage of all women. However, married young women who wished to delay the next pregnancies face several challenges in decision-making. For instance, the qualitative study found that women are often under pressure to have children once they get married due to traditional beliefs about fertility that prevent married women from using contraceptives. Women are expected to demonstrate their fertility after marriage, which may be even stronger for younger women as suggested by the literature,[33-37]. Moreover, in a social context such as Allada, where large families of seven children are the average, the family size is often decided by male partners and women feel less empowered to decide on their own to adopt contraceptives. This may explain the high total fertility rate of 5.7 in Benin. To get around the barrier of spousal consent, some married women request contraceptives discretely, after giving several births and as their households face economic challenges to raise the children. Most of the women would request long-term methods such as implants to be inserted in the leg or other unusual and less exposed parts of the body.

Our study revealed that factors associated with modern method use among those in need included literacy, not being married, having children, knowing a greater number of modern methods, and experiencing barriers to access health services. Although ‘literacy’ was not often used as a variable in previous studies, among young women there was evidence of a positive association between educational attainment and the demand satisfied,[34, 38]. Similarly, the number of modern methods known was positively associated with use. The strongest association was found between women’s marital status and the demand satisfied by a modern method. The multivariate analysis revealed that young unmarried women were four times more likely to use modern methods. This finding strengthens the evidence that married young women are being left behind in terms of demand satisfied. Also, a similar positive association between being unmarried and using modern contraceptives was found by Dansou et al. in a study among all women of reproductive age (15 to 49 years old) in Benin,[20]. Several other studies conducted in other sub-Saharan African countries such as Burkina Faso, Ethiopia, Nigeria, and Ghana found that modern method prevalence was higher among unmarried adolescent girls compared to married,[39, 40]. Young unmarried women used contraceptives to avoid pregnancy because of the social stigma associated with early pregnancy in Benin, and because they have no income to support a child on their own,[10, 20]. This finding is also supported by our qualitative data. Having a child was associated with modern method use. Other studies revealed that women with children were more likely to use modern methods,[41, 42]. Health education from care providers during pregnancy and in the postpartum period might be a contributing factor because the BDHS 2017-8 showed that coverage of antenatal care provided by skilled personnel and delivery in a health facility were 83% and 84%, respectively. Counterintuitively, we found that having barriers to access to health services was associated with a higher likelihood of modern methods use. We hypothesize that women who had accessed contraceptive methods were more likely to report barriers, while those who did not report such barriers did not theoretically exist.

Finally, our qualitative analysis revealed aspirin or paracetamol was used in Allada to prevent pregnancy, despite the lack of a biomedical evidence-based for such an effect. Boko et al. (2015) noted the same practice in Benin,[43]. The practice may be driven by the fact that requesting such medicine does not expose a young woman to any social stigma as the drug is not viewed as a contraceptive method by providers. This common practice may imply that the use of oral contraception use among young women in the DHS2017 -18 is an over-estimate. In a context such as in Allada where other ineffective drugs are used as oral contraceptives, it is important to promote accurate knowledge as well as practices through health education activities.

## Limitations

Our study has several limitations. We used secondary data, our analysis could have been affected by recall errors and biases in some variables collected on the DHS. For example, several assumptions were made in the DHS to assess the need for contraception among women, which were criticized to have possibly induced an underestimation in unmet needs for family planning,[44]. The assumptions concerned the frequency of sexual activity of women, women’s capacity to conceive (fecundity criteria, length of lactational amenorrhea which is 24 months in DHS versus 6 months clinically accepted standard), and the validity of their reports on whether recent births were desired. In the qualitative study, having a male interviewer might have affected responses from young women. Meanwhile, this was addressed as a young female interviewer continued interviews with female participants. Moreover, we could not ascertain whether a respondent in the qualitative study was sexually active whereas the analysis of determinants in the quantitative study focused on sexually active young women.

## Conclusion

Access to and use of modern contraceptive methods is low among women in Benin, especially among adolescent girls and young women. Adolescent girls and young women tend to use short-term methods such as condoms which they obtain from easily accessible providers and relatives. Unmarried and married women face different barriers to use modern contraceptives, which need to be carefully addressed. Most importantly young married women who wish to delay births should be specifically targeted in interventions to address challenges related to decision-making to use contraceptives.

Raising awareness in communities with strong culture towards encouraging early marriage and high fertility, as well as promoting comprehensive sexuality education and providing youth-friendly services may help reduce the incidence of unintended pregnancies among adolescent girls and young women,[45, 46].

## Data Availability

Quantitative data (BDHS 2017-18) is available from the DHS program (https://dhsprogram.com/). Qualitative data including interview audios and transcripts cannot be shared publicly because it contains information which enables one to identify each individual.

## Acknowledgments

We thank the Demographic Health Survey Program (http://dhsprogram.com/) for granting us access to the BDHS2017-18 dataset. We acknowledge TOTIN Mahoutondji Gilberte for her work as a research assistant during the qualitative data collection. We would like to thank the study participants for sharing their experiences as well as the local administrations of Allada for facilitating our study implementation.

## Contributors

NCAA, MM, MS, LB, TD, J-P D, and SG were involved in conception and study design. LB and MM provided statistical expertise. NCAA, AV, and CB collected qualitative data in the field. MM, MS, TD, CG, LK, and KP provided qualitative data analysis expertise. NCAA, MM, and MS were involved in the drafting of the manuscript. LB, TD, J-P D, and CG were involved in the critical revision of the manuscript for important intellectual content. All authors were involved in the f inal approval of the manuscript and the decision to submit the manuscript for publication.

## Patient and public involvement

Patients and/or the public were not involved in the design, conduct, reporting, or dissemination plans of this research.

## Patient consent for publication

Not required.

## Ethical statement

The BDHS 2017 -18 received approvals from the National Statistics Council and the National Ethics Committee for Health Research in Benin. As DHS data are secondary data, the quantitative strand did not require ethical approvals. The qualitative strand was conducted as part of the “ Continuum des Soins de Santé Reproductive au Bénin” study which received approvals from the Local Ethics Committee for Biomedical Research of the University of Parakou in Benin (approval number: 0092/CLERB-UP/P/SP/R/SA), the Institute of Tropical Medicine Antwerp in Belgium (approval number: IRB/AB/ AC/044), and Nagasaki University School of Tropical Medicine and Global Health in Japan (approval number: NU-TMGH_064). The participants in the interviews and the focus group discussions were informed of the research objectives, their voluntary participation in the study, and there was no disadvantage in refusal or withdrawal from the study. Written consent was obtained from each participant.

## References

1. United Nations Population Fund (UNFPA). Programme of Action. New York: UNFPA 1994.

2. Singh S, Sedgh G, Hussain R. Unintended pregnancy: worldwide levels, trends, and outcomes. Stud Fam Plann 2010;41(4):241–50. doi: 10.1111/j.1728-4465.2010.00250.x [published Online First: 2011/04/06]

3. Hubacher D, Mavranezouli I, McGinn E. Unintended pregnancy in sub-Saharan Africa: magnitude of the problem and potential role of contraceptive implants to alleviate it. Contraception 2008;78(1):73–8. doi: 10.1016/j.contraception.2008.03.002 [published Online First: 2008/06/17]

4. FP2020. Indicators 3-4 : Unmet Need and Demand Satisfied. http://2016-2017progressfamilyplanning2020org/en/measurement-section/unmet-need-and-demand-satisfied-indicators-3-4 [accessed November 2020]

5. Okigbo CC, Speizer IS. Determinants of Sexual Activity and Pregnancy among Unmarried Young Women in Urban Kenya: A Cross-Sectional Study. PLoS One 2015;10(6):e0129286. doi: 10.1371/journal.pone.0129286 [published Online First: 2015/06/06]

6. World Health Organization (WHO). Family planning evidence brief: family planning financing. Geneva: WHO, 2018. https://apps.who.int/iris/bitstream/handle/10665/255863/WHO-RHR-18.26-eng.pdf [accessed August 2019]

7. Darroch JE, Woog V, Bankole A, et al. Costs and Benefits of Meeting the Contraceptive Needs of Adolescents. New York: Guttmacher Institute 2016. http://www.guttmacher.org/sites/default/files/report_pdf/adding-it-up-adolescents-report.pdf [accessed May 2020]

8. Fescina R,B DM, Jl DR, et al. Guides for the PHC focused continuum of care of women and newborns. Montevideo: Latin American Center for Perinatology, Women and Reproductive Health, PAHO, 2009.

9. World Health Organization (WHO). Health for the world’s adolescents: A second chance in the second decade. Geneva: WHO, 2014

10. Sedgh G, Ashford LS, Hussain R. Unmet Need for Contraception in Developing Countries⍰: Examining Women’s Reasons for Not Using a Method. New York: Guttmacher Institute 2016. https://www.guttmacher.org/sites/default/files/report_pdf/unmet-need-for-contraception-in-developing-countries-report.pdf [accessed May 2020]

11. McCurdy RJ, Jiang X, Schnatz PF. Long-acting reversible contraception in adolescents in Sub-Saharan Africa: evidence from demographic and health surveys. Eur J Contracept Reprod Health Care 2018;23(5):357–64. doi: 10.1080/13625187.2018.1519535 [published Online First: 2018/11/23]

12. [dataset] Institut National de la Statistique et de l’Analyse Économique (INSAE) and ICF. Enquête Démographique et de Santé au Bénin, 2017-2018. Cotonou, Bénin and Rockville, Maryland, USA : INSAE and ICF. 2019.

13. Government of Benin. FAMILY PLANNING 2020 COMMITMENT. 2017. http://www.familyplanning2020.org/news/benin-fp2020-commitment [accessed March 2020]

14. Chae S, Woog V, Zinsou C, et al. Obstacles à la pratique contraceptive des femmes au Bénin. En Bref, New York: Guttmacher Institute 2015. https://www.guttmacher.org/sites/default/files/report_pdf/ib-benin-contraception-fr.pdf [accessed March 2020]

15. USAID, Communities AP. Benin’s Community-Based Access to Injectable Contraceptives Pilot Project. 2015. https://www.advancingpartners.org/sites/default/files/sites/default/files/resources/apc_benin_cba2i_pilot_brief_march_2015_english_final.pdf [accessed July 2020]

16. Okegbe T, Affo J, Djihoun F, et al. Introduction of Community-Based Provision of Subcutaneous Depot Medroxyprogesterone Acetate (DMPA-SC) in Benin: Programmatic Results. Glob Health Sci Pract 2019;7(2):228–39. doi: 10.9745/ghsp-d-19-00002 [published Online First: 2019/06/07]

17. Igras S, Diakité M, Lundgren R. Moving from theory to practice: A participatory social network mapping approach to address unmet need for family planning in Benin. Glob Public Health 2017;12(7):909–26. doi: 10.1080/17441692.2016.1147589 [published Online First: 2016/03/08]

18. Ahovey EC. Besoins non satisfaits en planification familiale au sein du couplelll: caractéristiques socio-démographiques et cadre de vie au Benin. 2002 <http://www.cicred.org/Eng/Seminars/Details/Seminars/Bangkok2002/001BangkokAhovey.pdf> [Accessed July 2020].

19. Sedgh G, Hussain R. Reasons for contraceptive nonuse among women having unmet need for contraception in developing countries. Stud Fam Plann 2014;45(2):151–69. doi: 10.1111/j.1728-4465.2014.00382.x [published Online First: 2014/06/17]

20. Dansou J, Adekunle AO, Arowojolu AO. Factors behind the preference in contraceptives use among non-pregnant and sexually active women in Benin Republic. Cent Afr J Public Health 2017;3(5):80–89. doi: 10.11648/j.cajph.20170305.15

21. MacQuarrie KLD. Unmet need for family planning among young women: levels and trends. DHS Comparative Reports No 34. Rockville, Maryland, USA: ICF International, 2014.

22. Radovich E, Dennis ML, Wong KLM, et al. Who Meets the Contraceptive Needs of Young Women in Sub-Saharan Africa? J Adolesc Health 2018;62(3):273–80. doi: 10.1016/j.jadohealth.2017.09.013 [published Online First: 2017/12/19]

23. Ahinkorah BO. Predictors of modern contraceptive use among adolescent girls and young women in sub-Saharan Africa: a mixed effects multilevel analysis of data from 29 demographic and health surveys. Contracept Reprod Med 2020;5(1):32. doi: 10.1186/s40834-020-00138-1.

24. Maxwell J. Designing a Qualitative Study. Qualitative Research Design An Interactive Approach (Third Edition) 2012

25. Hubacher D, Trussell J. A definition of modern contraceptive methods. Contraception 2015;92(5):420–1. doi: 10.1016/j.contraception.2015.08.008 [published Online First: 2015/08/16]

26. Bradley SEK, Croft T, Fishel J, et al. Revising Unmet Need for Family Planning. DHS Analytical Studies No 25 Calverton, Maryland, USA: ICF International 2012

27. Festin MP, Kiarie J, Solo J, et al. Moving towards the goals of FP2020 - classifying contraceptives. Contraception 2016;94(4):289–94. doi: 10.1016/j.contraception.2016.05.015 [published Online First: 2016/06/12]

28. Vigan A, Boyi C, Ahissou C, et al. Déterminants de l’utilisation des services le long du continuum des Soins de Santé Reproductive au Bénin: une exploration qualitative. Rapport d’étude. Ministère de la Santé, Centre de Recherche en Reproduction Humaine et Démographie (CERRHUD), Institut de Médecine Tropicale (IMT), 2020

29. . Hausmann-Muela S, Muela Ribera J, Toomer E, et al. The PASS-model: a model for guiding health-seeking behavior and access to care research. Malaria Reports 2012;2:e3. doi: 10.4081/malaria.2012.e3

30. Ochako R, Mbondo M, Aloo S, et al. Barriers to modern contraceptive methods uptake among young women in Kenya: a qualitative study. BMC Public Health 2015;15:118. doi: 10.1186/s12889-015-1483-1 [published Online First: 2015/04/18]

31. Campbell OM, Benova L, Macleod D, et al. Who, What, Where: an analysis of private sector family planning provision in 57 low- and middle-income countries. Trop Med Int Health 2015;20(12):1639–56. doi: 10.1111/tmi.12597 [published Online First: 2015/09/29]

32. Ali MM, Cleland J. Long term trends in behaviour to protect against adverse reproductive and sexual health outcomes among young single African women. Reprod Health 2018;15(1):136. doi: 10.1186/s12978-018-0576-6 [published Online First: 2018/08/16]

33. Asiimwe JB, Ndugga P, Mushomi J. Socio-demographic factors associated with contraceptive use among young women in comparison with older women in Uganda. DHS Working Papers No 95, 2013.

34. Asiimwe JB, Ndugga P, Mushomi J, et al. Factors associated with modern contraceptive use among young and older women in Uganda; a comparative analysis. BMC Public Health 2014;14:926. doi: 10.1186/1471-2458-14-926 [published Online First: 2014/09/10]

35. Ngome E, Odimegwu C. The social context of adolescent women’s use of modern contraceptives in Zimbabwe: a multilevel analysis. Reprod Health 2014;11:64. doi: 10.1186/1742-4755-11-64 [published Online First: 2014/08/12]

36. Shahabuddin AS, Nöstlinger C, Delvaux T, et al. What Influences Adolescent Girls’ Decision-Making Regarding Contraceptive Methods Use and Childbearing? A Qualitative Exploratory Study in Rangpur District, Bangladesh. PLoS One 2016;11(6):e0157664. doi: 10.1371/journal.pone.0157664 [published Online First: 2016/06/24]

37. Mardi A, Ebadi A, Shahbazi S, et al. Factors influencing the use of contraceptives through the lens of teenage women: a qualitative study in Iran. BMC Public Health 2018;18(1):202. doi: 10.1186/s12889-018-5116-3 [published Online First: 2018/02/01]

38. Tamang L, Raynes-Greenow C, McGeechan K, et al. Factors associated with contraceptive use among sexually active Nepalese youths in the Kathmandu Valley. Contracept Reprod Med 2017;2:13. doi: 10.1186/s40834-017-0040-y [published Online First: 2017/12/05]

39. Hounton S, Barros AJ, Amouzou A, et al. Patterns and trends of contraceptive use among sexually active adolescents in Burkina Faso, Ethiopia, and Nigeria: evidence from cross-sectional studies. Glob Health Action 2015;8:29737. doi: 10.3402/gha.v8.29737 [published Online First: 2015/11/13]

40. Marrone G, Abdul-Rahman L, De Coninck Z, et al. Predictors of contraceptive use among female adolescents in Ghana. Afr J Reprod Health 2014;18(1):102–9. [published Online First: 2014/05/07]

41. de Vargas Nunes Coll C, Ewerling F, Hellwig F, et al. Contraception in adolescence: the influence of parity and marital status on contraceptive use in 73 low-and middle-income countries. Reprod Health 2019;16(1):21. doi: 10.1186/s12978-019-0686-9 [published Online First: 2019/02/23]

42. Ahinkorah BO. Predictors of unmet need for contraception among adolescent girls and young women in selected high fertility countries in sub-Saharan Africa: A multilevel mixed effects analysis. PLoS One 2020;15(8):e0236352. doi: 10.1371/journal.pone.0236352 [published Online First: 2020/08/08]

43. Boko I, Baxerres C, Ouattara F, et al. Interroger au Bénin les usages populaires d’un médicament abortif, le misoprostol. Rev med perinat 2017;9(1):20–24. doi: 10.1007/s12611-016-0388-2

44. Bradley SE, Casterline JB. Understanding unmet need: history, theory, and measurement. Stud Fam Plann 2014;45(2):123–50. doi: 10.1111/j.1728-4465.2014.00381.x [published Online First: 2014/06/17]

45. International Planned Parenthood Federation (IPPF). Provide: Strengthening youth friendly services. London: IPPF, 2008.

46. UNESCO. UN urges Comprehensive Approach to Sexuality Education. <https://en.unesco.org/news/urges-comprehensive-approach-sexuality-education> [Accessed November 2020].

